# Accurate deep-learning model to differentiate dementia severity and diagnosis using a portable electroencephalography device

**DOI:** 10.1101/2024.11.01.24316610

**Authors:** Masahiro Hata, Takufumi Yanagisawa, Yuki Miyazaki, Hisaki Omori, Atsuya Hirashima, Yuta Nakagawa, Mitsuhiro Eto, Kenji Yoshiyama, Hideki Kanemoto, Byambadorj Nyamradnaa, Shusuke Yoshimoto, Kotaro Ezure, Shun Takahashi, Manabu Ikeda

**Author notes:** Corresponding author: Masahiro Hata Department of Psychiatry, Osaka University Graduate School of Medicine. 2-2 Yamadaoka, Suita, Osaka 565-0871, Japan.

## Abstract

Mild cognitive impairment (MCI) and dementia present critical health challenges in aging populations, highlighting the need for prompt and accurate diagnostic methods. Current diagnostic approaches for dementia are constrained by limited access to specialists and the high cost or invasiveness of the diagnostic methods, necessitating the development of widely accessible, cost-effective, and noninvasive alternatives. Electroencephalography (EEG) is a promising, accessible, cost-effective, noninvasive biomarker of dementia. However, traditional EEG systems are limited by their lack of portability and need for skilled technicians. This study proposes a deep-learning-based model to differentiate healthy volunteers (HVs) from patients with dementia-related conditions using data from portable EEG devices. A dataset of 233 participants, including 119 HVs and 114 patients categorized by severity and clinical diagnosis, was analyzed. We developed a customized transformer-based model to analyze the EEG data transformed into frequency-domain features using a short-time Fourier transform. The model was evaluated using 10-fold cross-validation and a hold-out dataset. In cross-validation, the model achieved an area under the curve (AUC) of 0.872 and a balanced accuracy (bACC) of 80.8% in distinguishing HVs from patients. Subgroup analyses by severity and clinical diagnosis yielded AUCs of 0.812 to 0.898 and bACCs of 74.9 to 86.4%, respectively. Comparable performance was observed in the holdout dataset, except for small sample sizes. These findings highlight the potential of portable EEG devices and deep-learning models as practical tools for dementia screening.

## 1. Introduction

Dementia affects >55 million people worldwide, with associated costs estimated to be $1.3 trillion annually. The number of patients and related expenses are expected to continue increasing [1]. In Japan, as of 2020, approximately 6.02 million people have dementia, and the prevalence of mild cognitive impairment (MCI) among those aged ≥65 is 13%, affecting approximately 4.71 million individuals [2]. Establishing a regional medical care system for dementia is essential; however, the number of dementia specialists in Japan is limited to approximately 2,000 [3]. Similar problems have been reported in the United States [4], suggesting that this is a global issue. Consequently, in areas with a shortage of specialists, the early diagnosis of dementia is challenging, leading to delays in diagnosis and therapeutic intervention.

The early diagnosis of dementia at a mild stage allows pharmacological and non-pharmacological interventions, potentially slowing disease progression [5]. Additionally, practical support for future planning and the establishment of a living environment that maintains an individual’s quality of life can reduce the burden of caregiving [6].

In addition, the accurate differential diagnosis of dementia diseases is essential to optimize clinical care, for instance, by carefully avoiding medications with anticholinergic properties and promoting aggressive identification of therapeutic targets, such as orthostatic hypotension and constipation [7]. The differential diagnosis of dementia largely depends on its clinical features, neuropsychological profiles, and various biomarkers, including structural magnetic resonance imaging (MRI), single-photon emission computed tomography (SPECT), amyloid positron emission tomography (PET), cerebral spinal fluid (CSF) markers, meta-iodobenzylguanidine (MIBG) myocardial scintigraphy, dopamine transporter (DAT) uptake in the basal ganglia, and polysomnography (PSG)-based confirmation of rapid eye movement (REM) sleep without atonia [8]. However, these investigations can be invasive, costly, require specialized skills, and are typically conducted in a limited number of institutions equipped with cutting-edge technology [9].

Owing to the global increase in the number of individuals with dementia [1], a strong demand exists for the development of noninvasive, inexpensive, and widely accessible diagnostic methods across numerous facilities.

Electroencephalography (EEG) is a diagnostic method that addresses these conditions. EEG is widely available, noninvasive, and inexpensive, making it a valuable tool in clinical practice, especially when compared with the high-precision biomarkers mentioned earlier. As brain activity primarily involves neuronal electrical activity, EEG and other electrophysiological approaches are considered highly reflective of brain function, making EEG a promising biomarker in dementia management [10].

EEG has recently been used as a promising tool to screen and assist in the diagnosis of dementia [11], and neurophysiological findings associated with neurodegenerative diseases have accumulated [10]. Recent studies have reported that the application of advanced machine learning [12,13] and deep learning techniques [11,14] to clinical electroencephalography (EEG) data significantly improves the accuracy of dementia identification.

We previously reported that deep learning models applied to clinical EEG data could accurately distinguish between multiple dementia-related diseases [15]. Furthermore, we demonstrated that deep learning models trained on the clinical EEG data of patients with dementia at one institute could accurately identify patients with dementia at the early MCI level, as well as those from other facilities [16].

These results suggest that EEG could serve as an auxiliary technology in the differential diagnosis of dementia. However, conventional multichannel clinical EEG systems have several disadvantages: they are expensive, large, and heavy (weighing 100 kg); require operation by specialized medical personnel; and require accurate placement of electrodes and paste on the scalp, which can be physically burdensome to the participants. Additionally, participants must move to the hospital’s examination room for EEG measurement, which involves logistical efforts and preparation time for the technician, posing barriers to screening in a large patient population.

Broader adoption of portable EEG devices is expected to address the limitations of clinical EEGs. For example, our previous research demonstrated that data obtained from portable EEG devices analyzed using deep learning techniques could effectively differentiate between patients with Alzheimer’s disease (AD) and healthy volunteers, albeit with a relatively small sample size [17]. Building on these preliminary findings, the present study aimed to develop a deep learning model to distinguish healthy volunteers and patients with dementia-related conditions using EEG data from a larger sample size of portable devices. Given the global increase in dementia patients, this approach could serve as a non-invasive and accessible screening tool for the early identification and classification of dementia-related conditions, potentially offering significant clinical benefits.

## 2. Methods

### 2.1 Study population

Portable EEG data were obtained from 294 participants (122 healthy volunteers (HVs) and 172 patients) at the Osaka University Hospital.

The HVs consisted of community-dwelling older adults. Their inclusion criteria were as follows: 1) no history of neurological or psychiatric diseases, 2) no history of severe head injury or alcohol/drug abuse, and 3) no impairment of daily living or global cognitive impairment (Mini-Mental State Examination (MMSE) [18] score ≥ 27) as in our previous study [15].

All patients underwent baseline assessments, including demographic, cognitive, and neuropsychiatric assessments; brain structure assessments using MRI or computed tomography; and laboratory measurements (e.g., blood cell count, blood chemistry measurements, thyroid hormone levels, vitamin B1, B12, and folic acid). The recruitment period was from April 2021 to February 2024. The following examinations were optionally performed according to the attending physician’s judgement: SPECT, CSF markers, MIBG myocardial scintigraphy, DAT uptake in the basal ganglia, and PSG. Based on these examinations, the patients were evaluated through expert conferences to determine their clinical diagnoses based on international criteria, and subsequent treatment plans. As in our previous study [15], this study included three dementia-related diseases: AD, Lewy body disease (LBD), and idiopathic normal-pressure hydrocephalus (iNPH). The patient details are described below.

Patients with AD were diagnosed according to international criteria [19,20], those with LBD based on standard diagnostic consensus criteria [8,21], and those with iNPH also met standard criteria [22]. The diagnoses of LBD and iNPH were prioritized over that of AD in patients considered to have LBD or iNPH with comorbid AD [19]. In addition, as a consequence of the expert conference, patients in whom a specific neurodegenerative or dementia-related disease could not be identified as the underlying pathology were included as “non-specific.”

Regarding the cognitive function of the patients’group, this study included those with MCI and dementia. Dementia severity was assessed based on the clinical dementia rating (CDR) [23]. In subsequent analyses, we considered MCI to be a CDR of 0.5 according to a previous study [24]; mild dementia to be a CDR of 1 [23]; and moderate dementia to be a CDR of 2 [23].

Under these conditions, the final analysis included 119 HVs and 114 patients. The 114 patients included 45 with MCI, 48 with mild dementia, and 21 with moderate dementia when categorized by severity, or 53 with AD, 32 with LBD, 22 with iNPH, and 7 with non-specific MCI when categorized by clinical diagnosis. The demographic and clinical information of the HVs and patients are summarized by severity in Table 1 and by clinical diagnosis in Table 2.

**Table 1.** Demographic and clinical data of the participants categorized by severity.

| CDR class | # of<br>Participa<br>nts | Sex<br>(male/female) | Age<br>(years±SD) | MMSE<br>(scores±SD) | Disease<br>(# of Participants) |
| --- | --- | --- | --- | --- | --- |
| HVs | 119 | 39/80 | 66.71±8.03 | 29.77±0.51 | - |
| MCI | 45 | 17/28 | 74.31±10.16 | 25.69±3.87 | LBD(15), AD(13), iNPH(10),<br>non-specific MCI(7) |
| Mild dementia | 48 | 19/29 | 75.46±9.33 | 20.52±3.61 | AD(28), LBD(13), iNPH(7) |
| Moderate dementia | 21 | 6/15 | 75.52±9.33 | 14.67±6.60 | AD(12), iNPH(5), LBD(4) |

MCI: Patients with mild cognitive impairrment
AD: Patients with Alzheimer's disease
LBD: Patients with Lewy body disease
iNPH: Patients with idiopathic normal pressure hydrocephalus
HV: Healthy volunteers
MMSE: Mini Mental State Examination
SD: Standard deviation

**Table 2.**
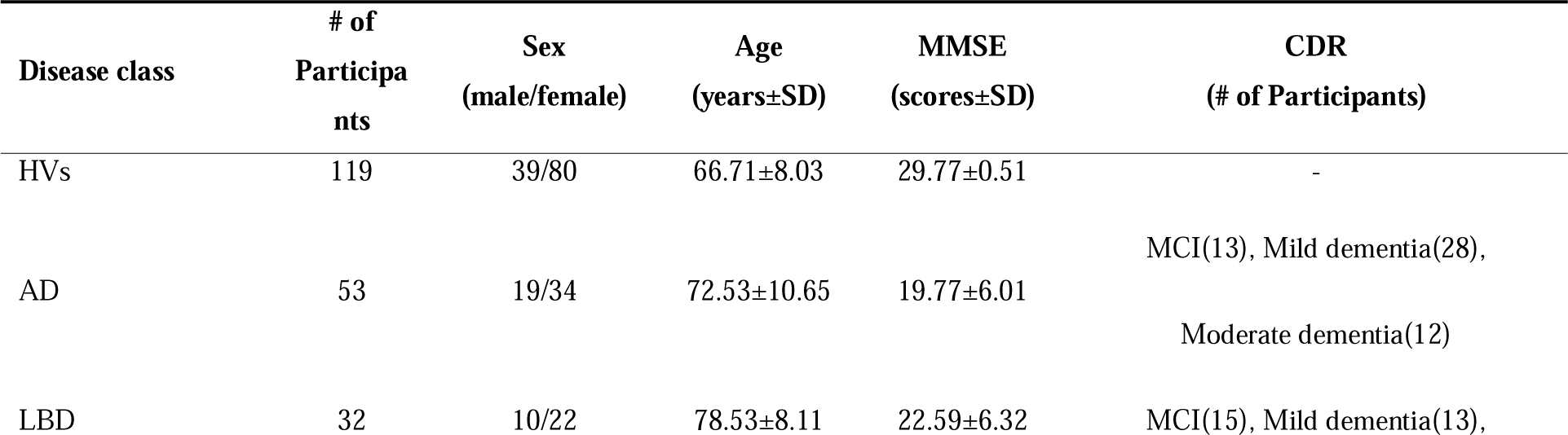
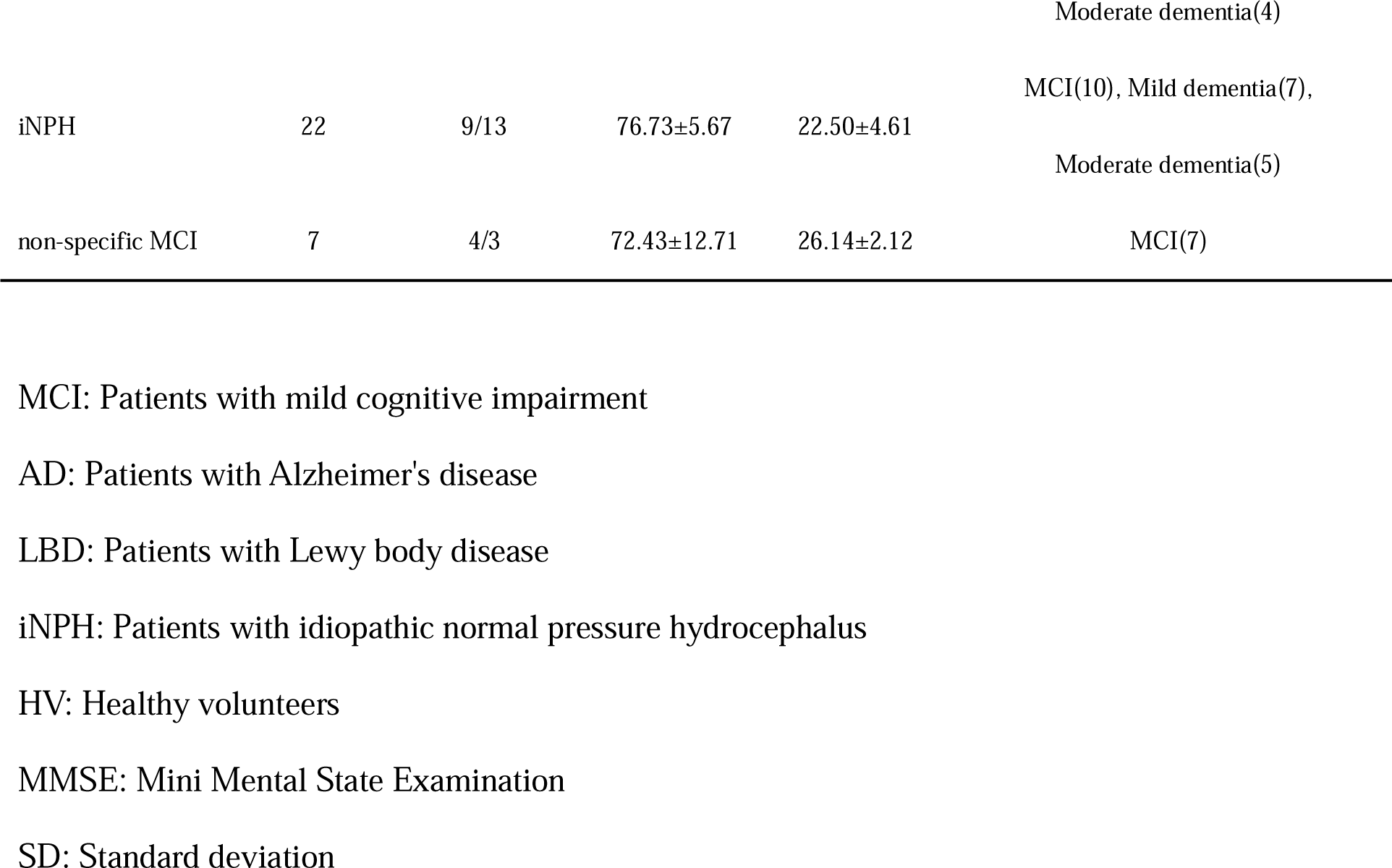
Demographic and clinical data of the participants categorized by clinical diagnosis.

### 2.2 Ethics

This study was performed according to the principles of the Declaration of Helsinki, approved by the Ethics Review Committee of Osaka University Hospital, and registered in the UMIN Clinical Trial Registry (UMIN 000042903). Prior to enrollment, we explained the use of their clinical data for this research to all participants and obtained their written informed consent. Each participant provided written informed consent after receiving a detailed explanation of the research procedures.

### 2.3 A patch-type portable EEG sensor

EEG measurements were conducted using a portable, patch-type EEG device, HARU-1 (Figure 1; PGV Inc., Tokyo, Japan). The HARU-1 has received medical approval from Japan’s Pharmaceuticals and Medical Devices Agency (PMDA) and has been evaluated using the same standards as traditional clinical EEGs (Certification Number: 302 AFBZX00079000, class II). EEG signals were measured using three channels, the ChZ (center), ChR (right), and ChL (left), with a sampling frequency of 250 Hz. The specifications of the wireless sensing device and the electrode sheet (Figure 1) are listed in Tables 3 and 4, respectively.

**Figure 1.**
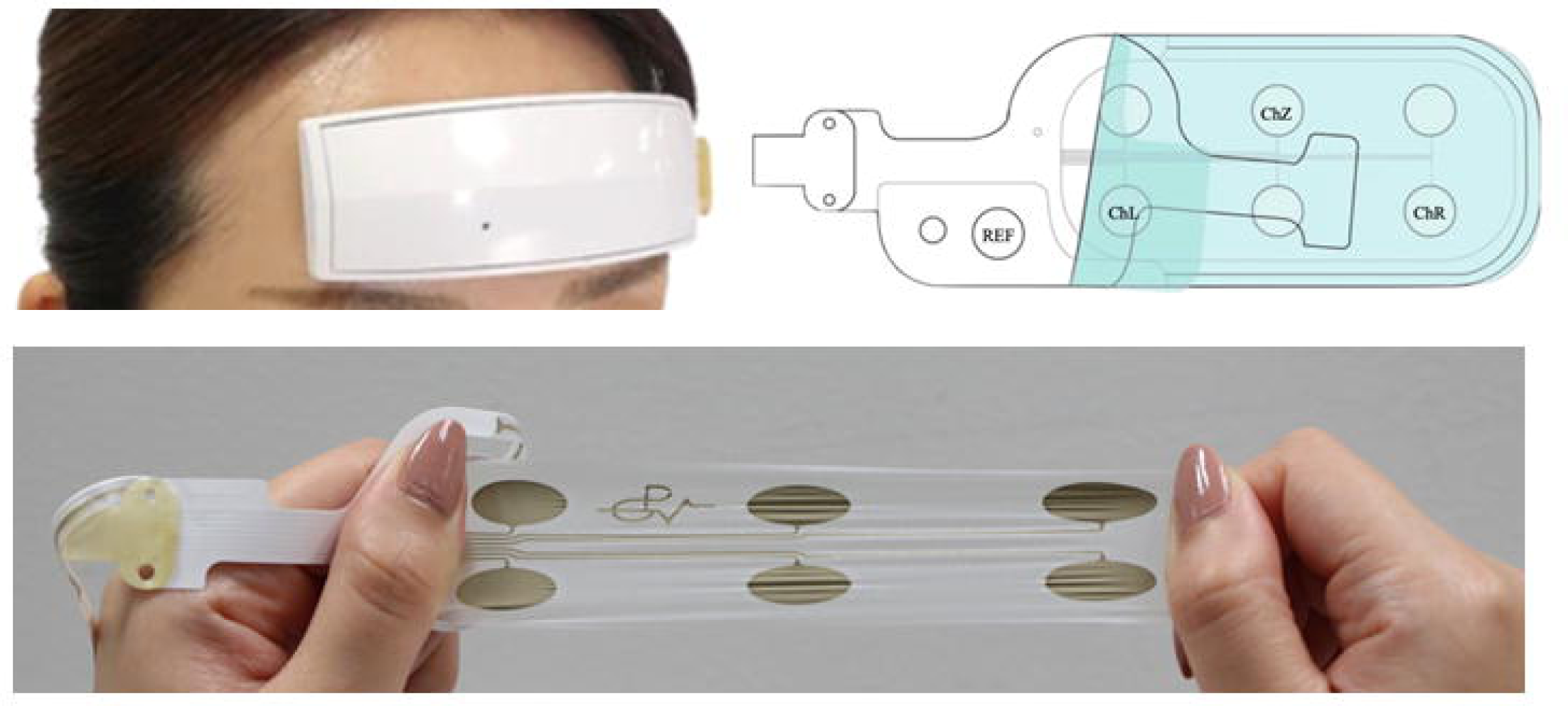
Patch-type EEG device and a disposable sheet with electrodes. EEG device (HARU) and electrode sheet in the worn state (top left). The sheet is connected to the device using a plug connector. The Haru sensor can be easily attached to the user’s forehead without any binding method. Stretchable electrode sheet (bottom). Positions of electrodes (top right). The 3-channel EEG (ChZ: center, ChR: right, ChL: left) were positioned on the forehead, while the reference electrode was designed to be attached to the mastoid process with its conductor cable detached from the sheet.

**Table 3.**
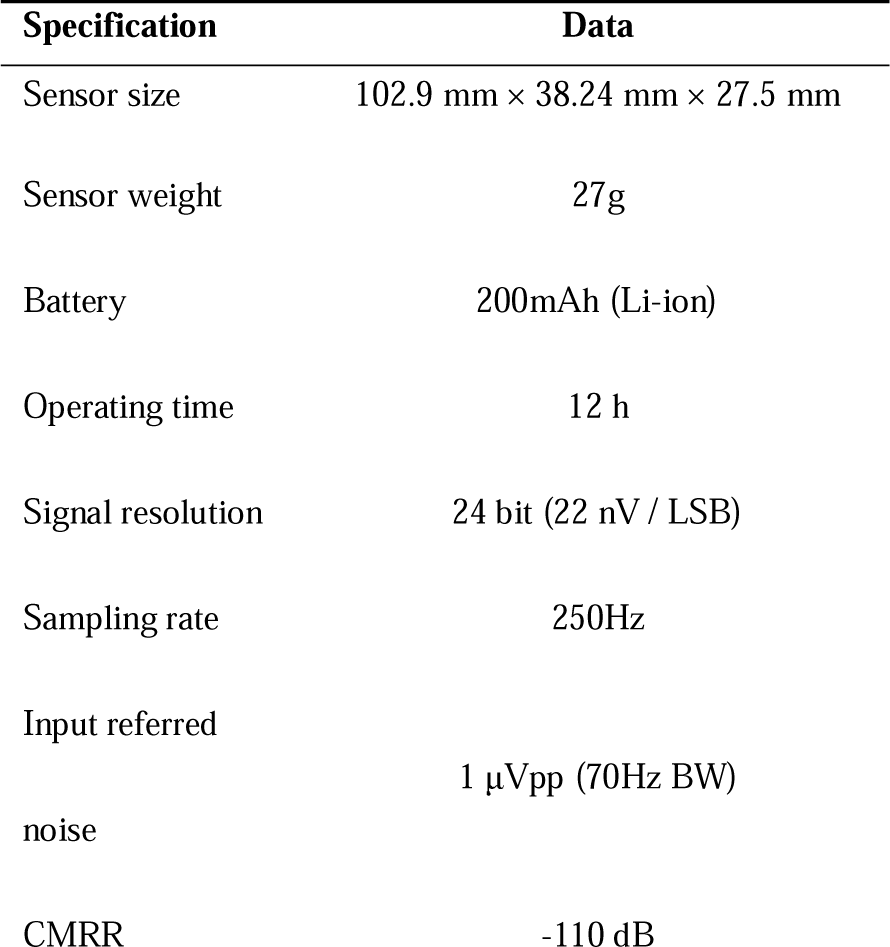
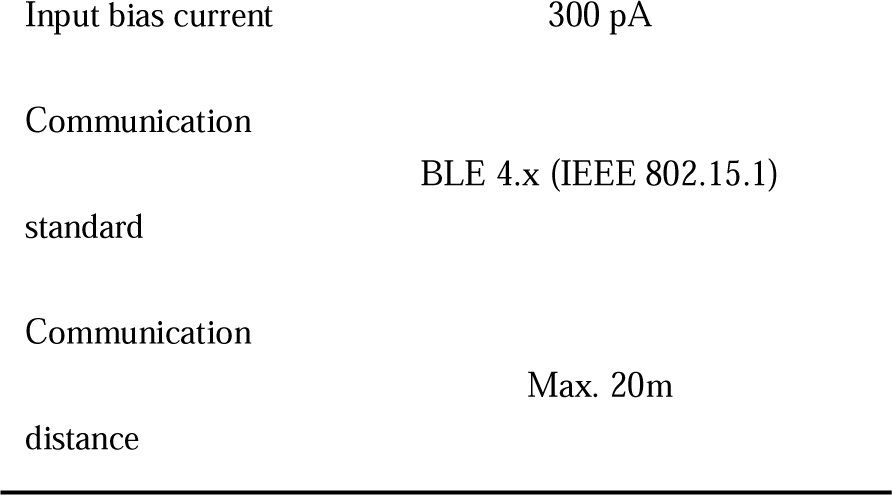
Specifications of the HARU-1 device.

**Table 4.** Sheet Specifications.

| Specification | Data |
| --- | --- |
| Sheet size | 50mm × 155mm × 50 μm |
| Sheet weight | Less than 1.0g |
| Moisture permeability | 2700 g/m<sup>2</sup>/day |
| Stretchability | 200% |

The device is lightweight, weighing 27 g, and has a curved shape designed to fit the user’s forehead comfortably. Its Li-ion battery, with a capacity of 200 mAh, is chargeable via a micro-USB connector and lasts for approximately 12 h in recording mode. The wireless communication interface is based on the Bluetooth Low Energy (BLE) protocol, which provides easy device control. The HARU-1 device boasts a high voltage resolution of up to 24 bits (22 nV/LSB) and low input-referred noise of 1 μVpp. The disposable electrode sheets (Notification Number: 13B2X10421000001, class I) of the device have a thickness of <50 μm, stretchability of up to 200%, and a moisture permeability of 2700 g/m²/day. These sheets are manufactured using a screen-printing process with a biocompatible gel on an elastic base and a silver-based material. The biocompatibility of the conductive and nonconductive gels used in these electrode sheets was assessed in accordance with ISO 10993 standards for skin sensitization, irritation, and in vitro cytotoxicity.

### 2.4 EEG preprocessing

The analysis used resting-state EEG data with eyes closed. As part of the data preprocessing, a band-pass filter (0.5–95 Hz), notch filters (50 and 60 Hz), and cutting of the first 5 s were applied to reduce the power line noise and artifacts, thereby enhancing the signal quality. The filtered EEG data were subjected to short-term Fourier transform (STFT) to analyze the frequency content over time [25]. The STFT was computed using the following parameters: Hamming window, segment length (n_perseg_) of 8 s (2000 samples), overlap (n_overlap_) of 7 s (1750 samples), and FFT length (n_fft_) of 2048 points.

The STFT is defined as follows:

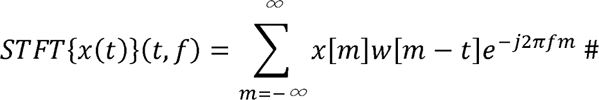

where *x(t)* represents the signal, *w(t)* is the Hamming window function, t is the time, and f is the frequency. The Hamming window helps minimize spectral leakage.

In this study, the segment length was set to 8 s, allowing for a detailed frequency analysis while maintaining an adequate time resolution. An overlap of 7 s ensured a high degree of temporal continuity between segments, further enhancing the resolution of transient events in the EEG data. An FFT length of 2048 points provided a high-resolution frequency spectrum, facilitating the detection of subtle changes in EEG signals.

After applying the STFT, the axes were rearranged and the absolute values of the amplitude spectra were obtained. As the frequency features are given by *n_fft_*/2 +1, this resulted in 1025 frequency bins. Finally, the 2-min EEG data for each participant were transformed into frequency features with dimensions of 108 × 1025 × 3, where 108 was the number of 8-s epochs, 1025 was the number of frequency bins, and 3 was the number of channels.

### 2.5 Data splitting

First, the dataset was divided into cross-validation and holdout datasets at a 0.9 to 0.1 ratio. As a result of the split, the holdout dataset included 12 HVs and 12 patients, whereas the cross-validation dataset included 107 HVs and 102 patients. Within the cross-validation dataset, the data was further split into training and validation sets in a 0.9 to 0.1 ratio. A 10-fold cross-validation method was employed, applying the Stratified K Fold [26] to ensure that the distribution of diseases and CDR was consistent across all folds, thus minimizing bias.

### 2.6 Model Architecture

A transformer-encoder-based model was deployed to perform the classification task. The architecture of the model is illustrated in Figure 2. The model architecture was based on a customized transformer encoder. However, several modifications were made to the standard transformer encoder [27], to facilitate the extraction of features from the 3-channel EEG data. The modifications were as follows:

**Figure 2.**
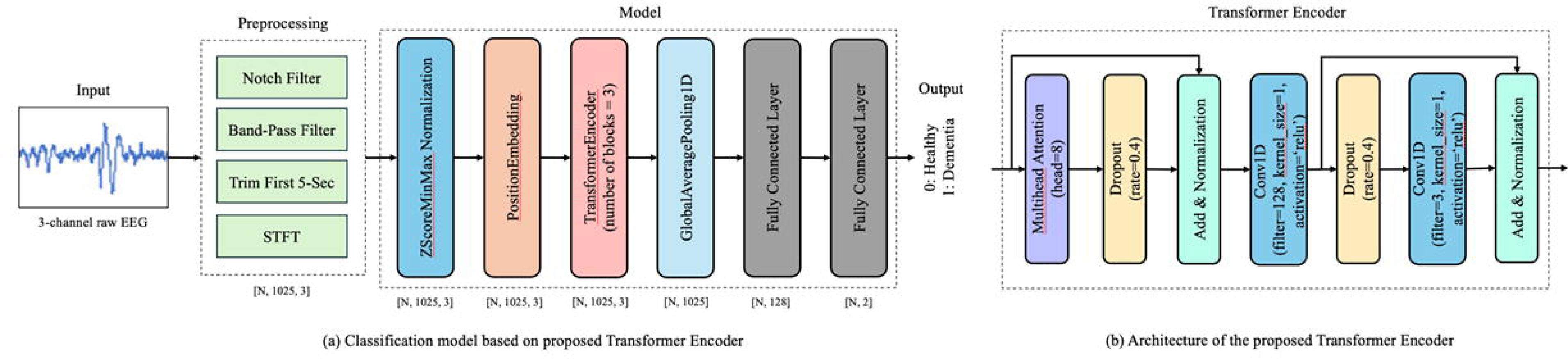
Model Architecture. The model architecture is based on a customized transformer encoder. (a) Classification model on proposed transformer encoder (b) Architecture of the proposed transformer encoder, where ‘N’ refers to the batch size.

First, instead of a single transformer encoder, three parallel transformer encoder blocks were used. This parallelization allowed the model to capture diverse features from the input data, thereby enhancing its robustness and accuracy. In addition, the traditional feedforward network was replaced by a convolutional feedforward network using Conv1D layers [28]. This change enhanced the model’s ability to capture local dependencies in the data, making it particularly useful for learning inter-channel relationships in time-series data. Furthermore, to prevent overfitting, enhanced dropout regularization [29] was applied at multiple stages within each transformer encoder block. This improved the generalization performance of the model on unseen data. Finally, as in the conventional architecture, layer normalization was applied after both the multi-head attention mechanism and feedforward network. This step was crucial for stabilizing the training process and ensuring a faster convergence. Using these customizations, the model was effectively adapted to extract features from 3-channel EEG data.

### 2.7 Model training

To evaluate the model, we conducted a 10-fold cross-validation, resulting in the creation of 10 models [30]. In each fold, 90% of the dataset was used for training and the remaining 10% was used for validation. An ADAM [31] optimizer with a learning rate of 0.0001 was used for each cross-validation. To prevent overfitting, an early stopping mechanism was introduced. Specifically, if no improvement in the validation metrics was observed for 50 consecutive epochs, training was halted and the best weights observed during training were restored [32].

In each fold, features were generated per participant, with 108 epochs of features created per participant. The epoch-wise features of the entire training dataset were concatenated, randomly shuffled to avoid order dependency during training, and fed into the model using a batch size of 32. This approach ensured the integrity and reliability of the data. To maintain consistency, the data were not shuffled during the calculation of the validation metrics.

The main libraries and their versions used in the training process were numpy [33] version 1.24.3, scipy [34] version 1.12.0, and TensorFlow [35] version 2.13.0.

Using this setup, the performance of the model was comprehensively evaluated, and its robustness and reliability were confirmed.

### 2.8 Model inference

The model outputs were generated per epoch, and the epoch-wise results were averaged for each participant to obtain participant-level results. In the evaluation of the 10-fold cross-validation dataset, the results for each participant in the validation dataset of each fold were aggregated to assess the overall performance. An ensemble method was adopted to evaluate the holdout dataset by averaging the outputs of the 10 models trained through cross-validation. Specifically, we averaged the epoch-wise results for each participant and further averaged the output results of each of the 10-fold models to obtain the results for the holdout participants.

### 2.9 Evaluation Metrics

For the binary classification task of distinguishing between the HVs and patients, we employed several evaluation metrics to comprehensively assess the performance of our model. The selected metrics included sensitivity, specificity, balanced accuracy (bACC), and area under the receiver operating characteristic (ROC) curve (AUC) [34,35]. bACC

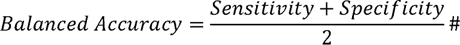

The bACC is the average of the sensitivity and specificity, providing a single metric that considers both types of classification errors. This is particularly useful for imbalanced datasets [38].

Using these evaluation metrics, we performed a comprehensive assessment of the ability of our model to distinguish between HVs and patients. Each metric provided valuable insights into the different aspects of model performance, contributing to a robust evaluation framework.

### 2.10 Statistical analyses

To compare the ages of the HVs and patients, one-way analysis of variance (ANOVA) was conducted across all groups. Post-hoc pairwise comparisons were subsequently performed using t-tests with Bonferroni correction. Sex distribution was assessed using the Fisher’s exact test. In the post-hoc analysis, assuming a two-tailed test with an effect size of 0.5, the power was calculated to be 96.8% for the sample sizes of HVs and patients. The significance level for all statistical tests was set at 0.05.

## 3. Results

### 3.1 Demographic and clinical data analysis

When comparing the HVs and patients categorized by severity, a significant difference (p = 4.19×10^-10^) in age was observed. Post-hoc analysis revealed significant differences between the HVs and each patient group (HVs vs. MCI: p = 2.74 ×10^-5^, HVs vs. mild dementia: p = 2.17×10^-7^, HVs vs. moderate dementia: p = 4.00×10^-4^, respectively), while no significant differences were detected among the patient groups themselves. When comparing the HVs and patients categorized by clinical diagnosis, a significant difference (p = 1.30×10^-11^) in age was observed. Post-hoc analysis revealed significant differences in age between the HVs and each patient group (HVs vs. AD: p = 6.51 ×10^-4^, HVs vs. LBD: p = 2.11×10^-9^, HVs vs. iNPH: p = 1.79×10^-8^), however, no significant difference was observed between the HVs and non-specific MCI.

In the comparison of sex distribution (male-to-female ratio) among HVs and patient groups according to severity and clinical diagnosis, no significant differences were revealed (p = .745 and p= .677, respectively).

### 3.2 Differentiating between HVs and patients using a deep-learning model

We achieved an AUC of 0.872 and bACC of 80.8% for distinguishing the HVs from the patients. Comparisons between the HVs and patients across severity levels resulted in an AUC of 0.862 and bACC of 79.1% ()HVs vs. MCI), AUC of 0.888 and bACC of 81.1% (HVs vs. mild dementia), and AUC of 0.858 and bACC of 83.7% (HVs vs. moderate dementia). (Table 5). These results are summarized in the receiver operating characteristic (ROC) curves for the four classifications (Figure 3).

**Table 5.**
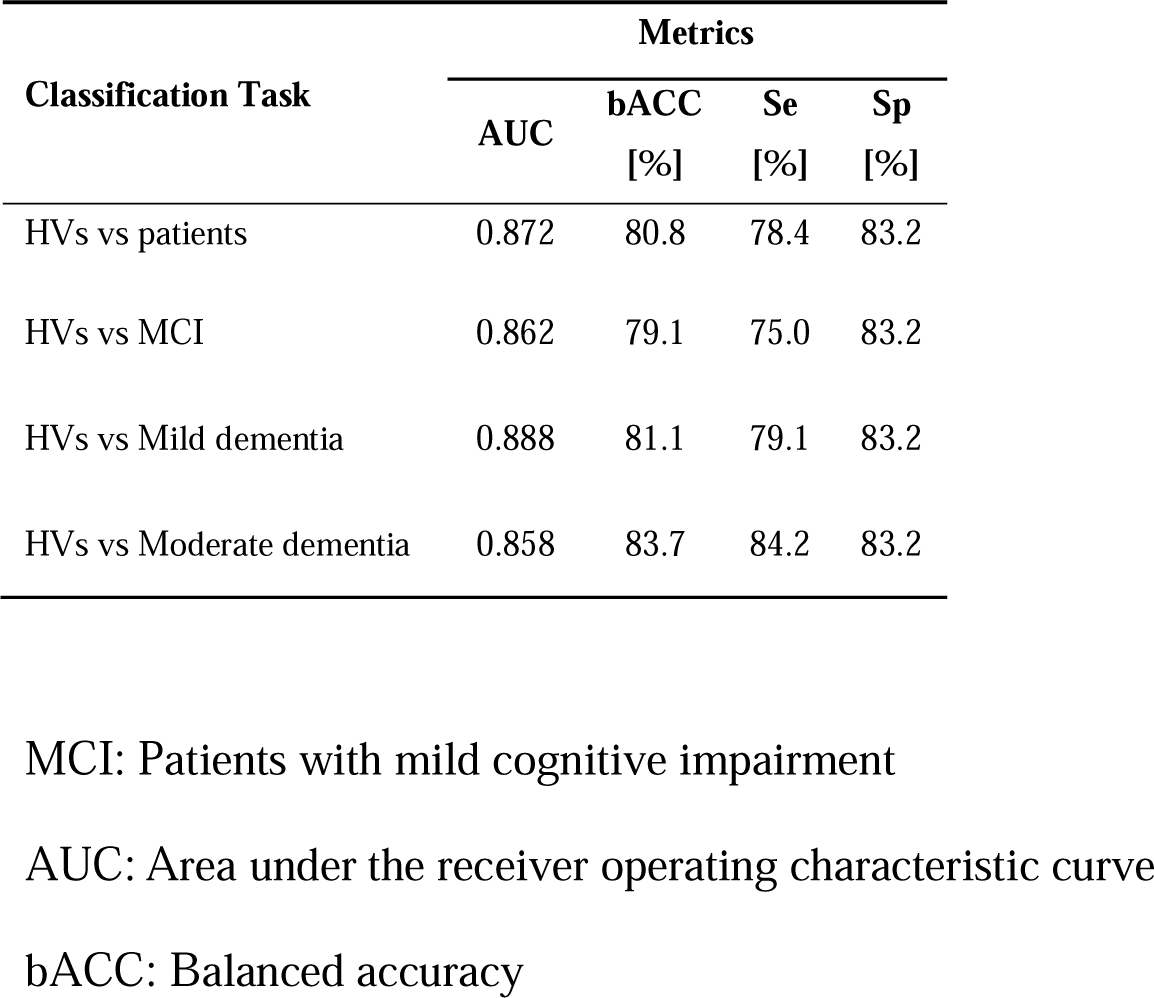
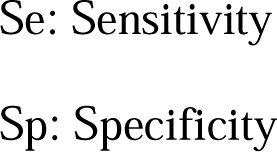
Results of the 10-fold cross validation method categorized by severity.

| Classification Task | Metrics |  |  |  |
| --- | --- | --- | --- | --- |
|  | AUC | bACC [%] | Se [%] | Sp [%] |
| HVs vs patients | 0.872 | 80.8 | 78.4 | 83.2 |
| HVs vs MCI | 0.862 | 79.1 | 75.0 | 83.2 |
| HVs vs Mild dementia | 0.888 | 81.1 | 79.1 | 83.2 |
| HVs vs Moderate dementia | 0.858 | 83.7 | 84.2 | 83.2 |

MCI: Patients with mild cognitive impairment
AUC: Area under the receiver operating characteristic curve
bACC: Balanced accuracy
Se: Sensitivity
Sp: Specificity

**Figure 3.**
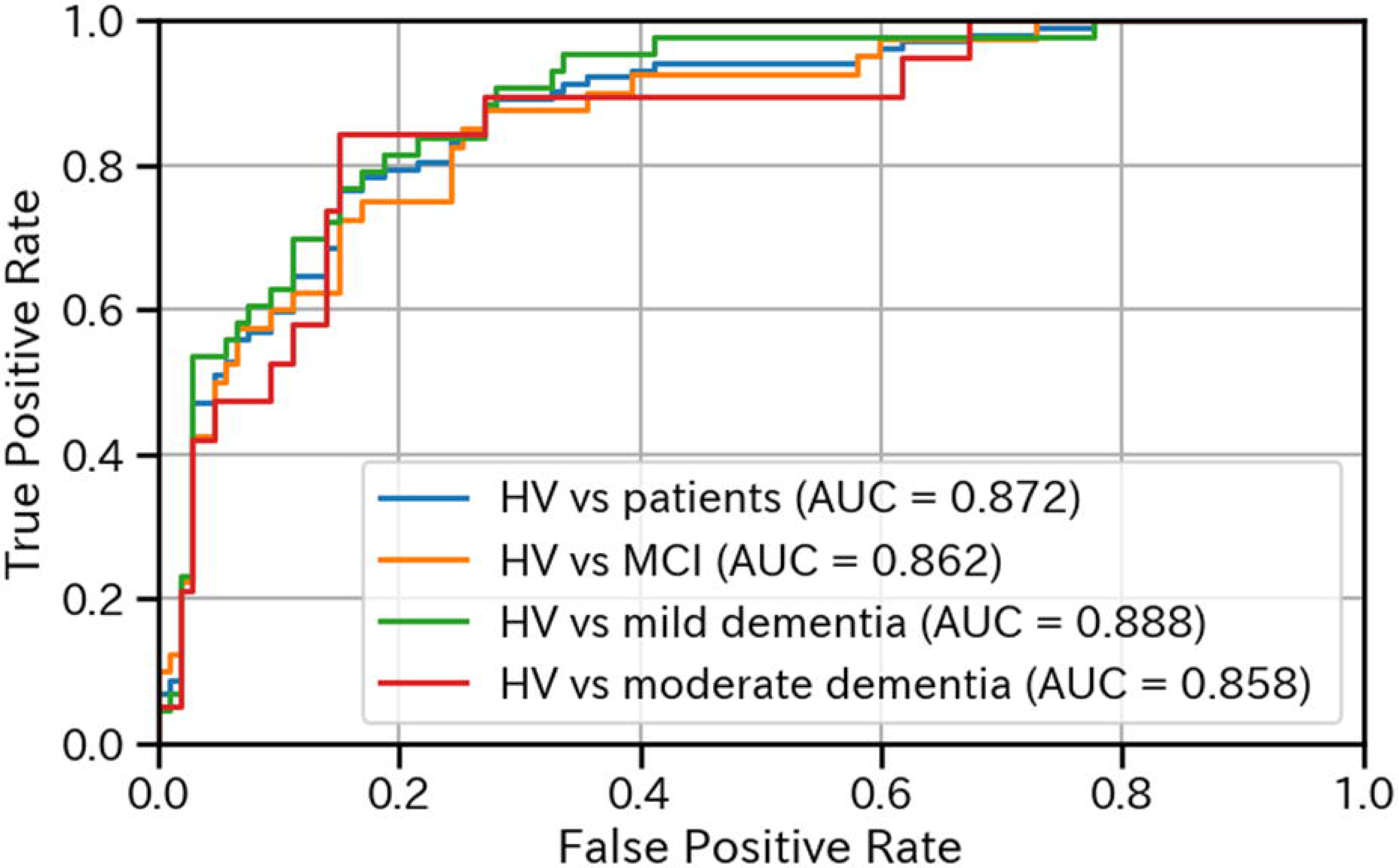
Receiver operating characteristic curve for classifications based on severity. HV: Healthy volunteers MCI: Patients with mild cognitive impairment AUC: Area under the receiver operating characteristic curve

We also conducted a sub-analysis to assess the impact of age on the differentiation process. Given that the number of HVs was larger than that of each subgroup (MCI, mild dementia, and moderate dementia), we adjusted the number of HVs to match that of each subgroup. The adjustment method involved random selection of the same number of HVs as those in each subgroup. This random selection was repeated until no significant age difference was observed between each subgroup and selected HVs. Consequently, for the HVs vs. MCI the bACC was 78.7% (79.1% before adjustment), for the HVs vs. mild dementia it was 76.7% (81.1% before adjustment), and for the HVs vs. moderate dementia it was 84.2% (83.7% before adjustment), with an average decrease in bACC of 1.4% after adjustment.

Additionally, comparisons between the HVs and patients classified by clinical diagnosis yielded an AUC of 0.866 and bACC of 77.8% (HVs vs. AD), AUC of 0.898 and bACC of 86.4% (HVs vs. LBD), AUC of 0.867 and bACC of 81.6% (HVs vs. iNPH), and an AUC of 0.812 and bACC of 74.9% (HVs vs. nonspecific MCI) (Table 6). These results are summarized in the ROC curves of the four classifications (Figure 4).

**Figure 4.**
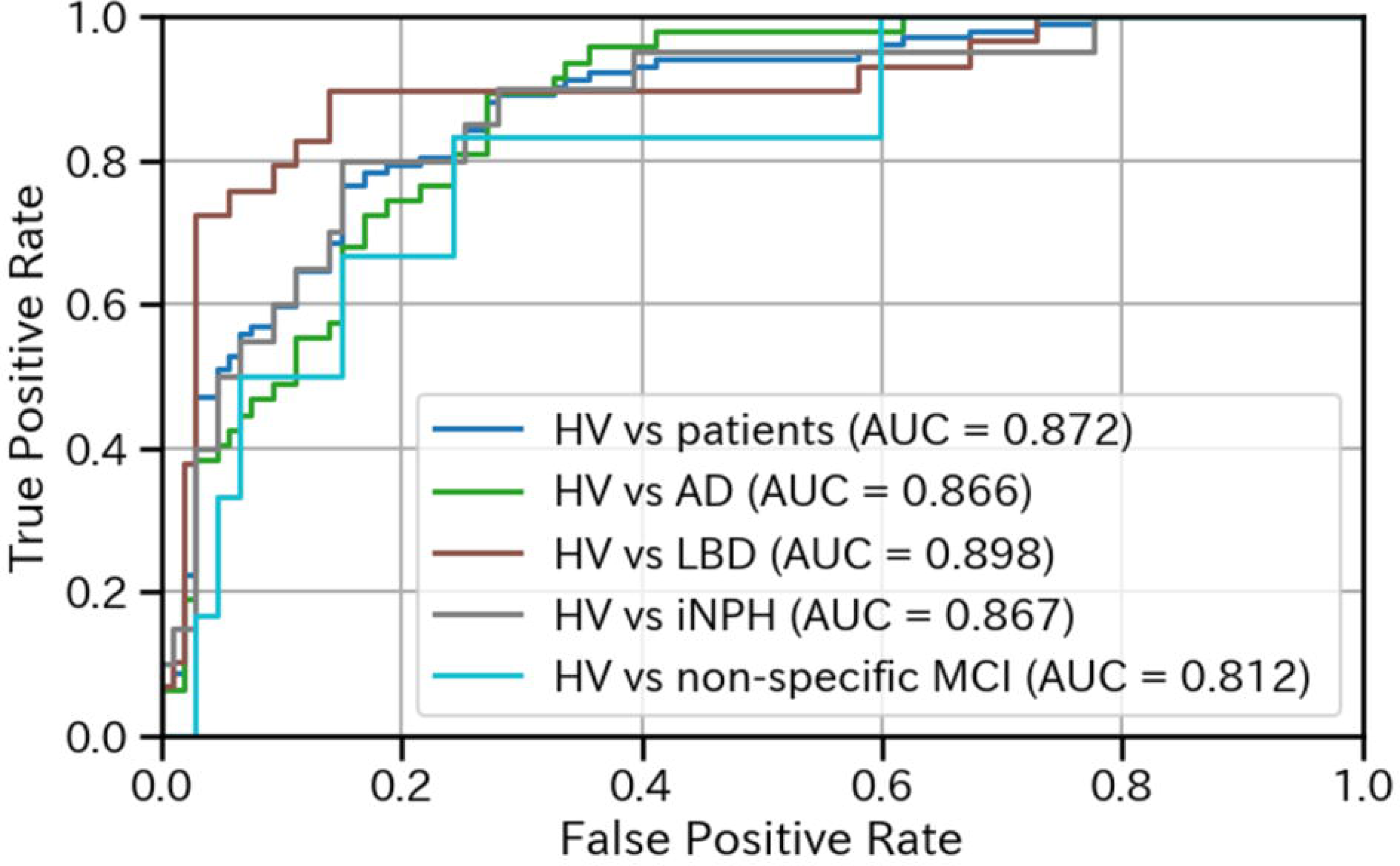
Receiver operating characteristic curve for classifications based on clinical diagnosis. HV: Healthy volunteers AUC: Area under the receiver operating characteristic curve AD: Patients with Alzheimer’s disease LBD: Patients with Lewy body disease iNPH: Patients with idiopathic normal pressure hydrocephalus MCI: Patients with mild cognitive impairment

**Table 6.** Results of the 10-fold cross validation method categorized by clinical diagnosis.

| Classification Task | Metrics |  |  |  |
| --- | --- | --- | --- | --- |
|  | AUC | bACC<br>[%] | Se<br>[%] | Sp<br>[%] |
| HVs vs patients | 0.872 | 80.8 | 78.4 | 83.2 |
| HVs vs AD | 0.866 | 77.8 | 72.3 | 83.2 |
| HVs vs LBD | 0.898 | 86.4 | 89.7 | 83.2 |
| HVs vs iNPH | 0.867 | 81.6 | 80.0 | 83.2 |
| HVs vs non-specific MCI | 0.812 | 74.9 | 66.7 | 83.2 |

AUC: Area under the receiver operating characteristic curve
bACC: Balanced accuracy
Se: Sensitivity
Sp: Specificity
HV: Healthy volunteers
AD: Patients with Alzheimer's disease
LBD: Patients with Lewy body disease
iNPH: Patients with idiopathic normal pressure hydrocephalus
MCI: Patients with mild cognitive impairment

We then examined the impact of age on the classification, using the same method as that used for the analysis based on the severity of dementia. Consequently, for the HVs vs. AD the bACC was 80.8% (77.8% before adjustment), for the HVs vs. LBD it was 79.3% (86.4% before adjustment), for the HVs vs. iNPH it was 75.0% (81.6% before adjustment), and for the HVs vs. nonspecific MCI it was 83.3% (74.9% before adjustment), with an average increase in bACC of 0.5% after adjustment.

The holdout analysis demonstrated an AUC of 0.882 and bACC of 87.5% between the HVs and patients (Table 7). Sensitivities in the holdout set based on clinical diagnosis were 83.3% for patients with AD, 100.0% for patients with LBD, 50.0% for patients with iNPH, and 0.0% for nonspecific MCI, noting that the latter result was based on only one participant (Table 8).

**Table 7.** Results of the Holdout dataset categorized by severity.

| Classification Task | Metrics |  |  |  |
| --- | --- | --- | --- | --- |
|  | AUC | bACC<br>[%] | Se<br>[%] | Sp<br>[%] |
| HVs vs patients | 0.882 | 87.5 | 83.3 | 91.7 |
| HVs vs MCI | 0.817 | 85.8 | 80.0 | 91.7 |
| HVs vs Mild dementia | 0.900 | 85.8 | 80.0 | 91.7 |
| HVs vs Moderate dementia | 1.00 | 95.8 | 100 | 91.7 |

MCI: Patients with mild cognitive impairment
AUC: Area under the receiver operating characteristic curve
bACC: Balanced accuracy
Se: Sensitivity
Sp: Specificity

**Table 8.**
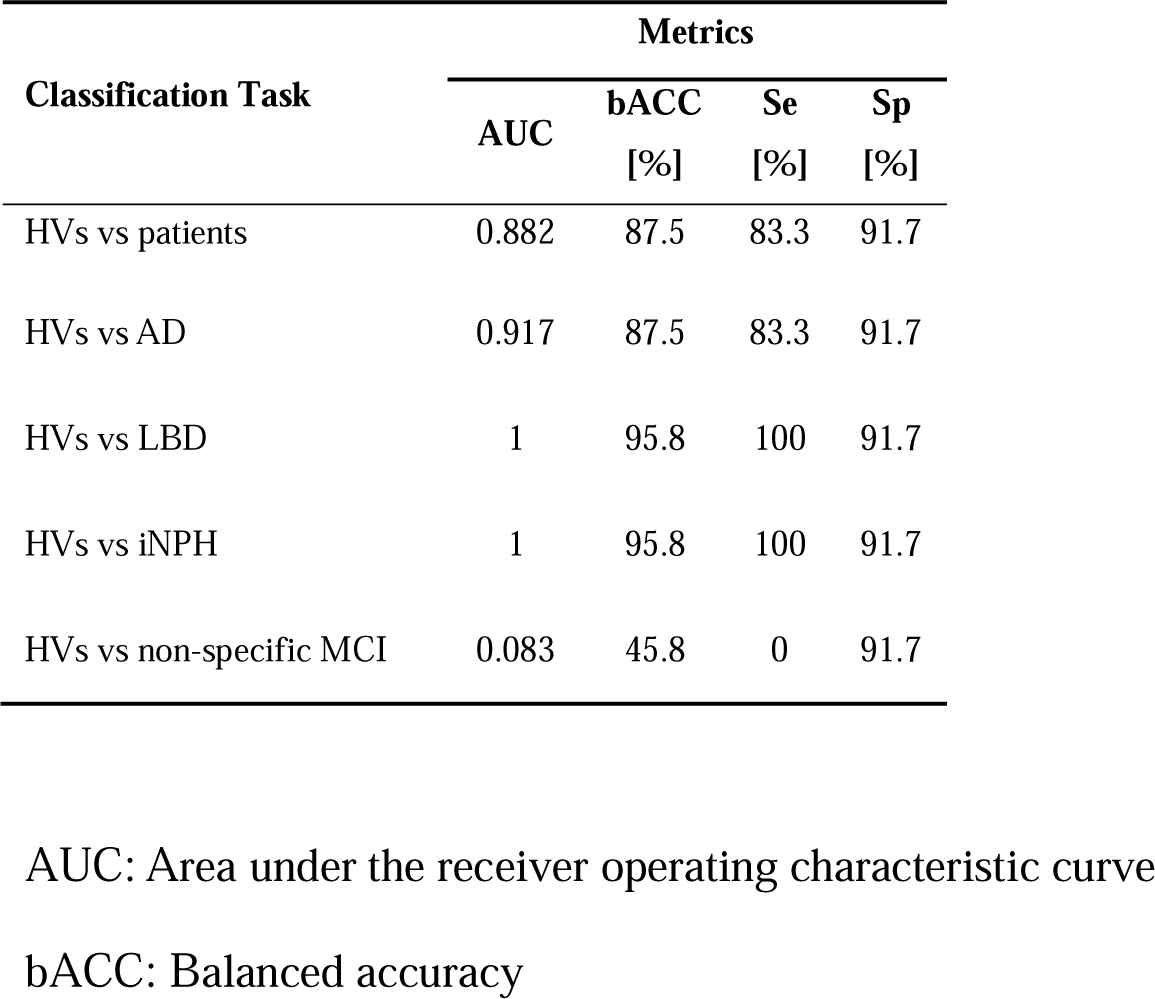
Result of the Holdout dataset categorized by clinical diagnosis.

## 4. Discussion

In this study, we developed a deep-learning model that uses data from a portable EEG device to differentiate HVs and patients with dementia-related conditions. Our cross-validation analysis yielded an AUC of 0.872 and a bACC of 80.8% for distinguishing HVs from patients. Severity-based comparisons produced AUCs ranging from 0.858–0.888 and bACCs between 79.1% and 83.7%, whereas clinical diagnosis-based comparisons produced AUCs of 0.812–0.898 and bACCs ranging from 74.9–86.4%. These results confirm that a high classification accuracy was achieved in all identification tasks.

A previous study, which applied deep learning to EEG data from 19 channels [39], reported an AUC of 0.86 and an accuracy of 0.79 for distinguishing HVs from patients with mild cognitive impairment (MCI) or dementia, including multiple clinical diagnoses as in our study. While their results are comparable with ours, our model demonstrated slightly superior performance despite using only a portable 3-channel EEG device. This highlights the practicality of the proposed approach, which offers competitive accuracy with fewer channels.

Additionally, our results indicated that classification accuracy, as measured by the bACC, increased with dementia severity. This trend is consistent with the findings of Fan et al. [40], who applied machine learning to 19-channel EEG data to differentiate HVs from patients with AD based on CDR scores. However, their study was limited to patients with AD, whereas our study encompassed a broader range of clinical diagnoses. In Fan et al.’s study, the accuracy ranged from 42–56% for HVs vs. MCI, 59–70% for HVs vs. mild dementia, and 58–79% for HVs vs. moderate dementia. Notably, in our study, even in distinguishing MCI, where Fan et al. [40] reported lower accuracy, we achieved higher precision, suggesting that our model is better at detecting early-stage dementia.

When comparing our findings with those of other studies focused on MCI vs. HV classification, deep learning models applied to 19-channel EEG data have shown accuracies ranging from 75% [41] to as high as 92% [11,14]. Although the accuracy of our model (79%) exceeds that of the study reported by Kim et al. [41], it remains lower than that in higher-performing models [11,14]. However, our model used only three frontal channels, which offers significant practical advantages in terms of accessibility and feasibility for widespread use.

In the results based on clinical diagnosis, previous studies employing machine learning and deep learning algorithms for EEG-based AD classification have reported varying degrees of success, with accuracies ranging from 80–95% [42–44]. However, these studies were limited by relatively small sample sizes, which may have constrained the generalizability and robustness of their results. Even in a study with >100 participants, which secured a substantial number of patients and compared AD and HV classifications using machine learning [13], that achieved an AUC of 0.76, our model demonstrated higher accuracy, indicating that our approach provides superior performance despite using fewer EEG channels. In comparison to Fan et al. [40], who reported an accuracy in the high 60% range for distinguishing HVs from patients with AD, even when averaging the best results across multiple models for different disease severity levels, our study demonstrated superior performance, achieving approximately 10% higher accuracy in this classification task. However, when compared with studies such as those by Morabito et al. [14] and Ieracitano et al. [11], which reported accuracies of 85% and 91%, respectively, our model lags behind in terms of accuracy. This discrepancy may be attributed to the reduced information available from the portable 3-channel EEG used in our study, which could limit further improvements in classification accuracy.

Limited research has been conducted on the application of machine-learning or deep-learning techniques to EEG data in patients with LBD. One exception was the study by Dauwan et al. [12], which applied machine learning to clinical EEG data and achieved a discrimination rate of approximately 90% between HVs and patients with LBD. Our findings are consistent with this, as the highest classification accuracy in our study was observed in distinguishing LBD, likely because of the presence of distinct neurophysiological disturbances commonly associated with LBD, as outlined in the international clinical diagnostic criteria [8]. These disturbances could be captured effectively by our deep learning model, highlighting its ability to identify characteristic patterns specific to LBD.

To our knowledge, no prior studies, aside from our own, have applied machine learning or deep learning to EEG data from patients with idiopathic normal pressure hydrocephalus (iNPH). Moreover, our previous work remains the only study attempting to distinguish between AD, LBD, and iNPH using EEG data. Compared with our earlier studies [15,16], the classification accuracy in this study was 4–15% lower across clinical diagnoses, likely because of the limited number of EEG channels used in this study. This suggests that although our model has made advancements, the reduction in available data when using a portable EEG device presents challenges in achieving the same level of accuracy as that achieved with models using 19-channel clinical EEG systems.

This study had a few limitations. First, we extracted only specific groups with defined clinical diagnoses from consecutively measured portable EEG cases collected during the study period. Consequently, there was some variability in both the cognitive function levels and sample sizes for each clinical diagnosis. Therefore, these results should be interpreted with caution. Second, with respect to the age gap between the HVs and patient groups, the bACC in distinguishing between HV and patients with each severity level of dementia and each clinical diagnosis fluctuated by no more than 2% in the analysis adjusted for age differences. Thus, the influence of age on discrimination was minimal, indicating that factors other than age might have played a significant role in the observed discrimination.

## Conclusions

In this study, we developed a deep-learning model to differentiate HVs and patients with dementia-related conditions using a patch-type portable EEG device. Our model demonstrated high accuracy in distinguishing HVs from patients as well as in classifying dementia according to severity and clinical diagnosis. Using the mobility of a portable EEG device, the model holds promise for clinical use, particularly in facilitating early detection and preliminary evaluation in a variety of clinical settings.

## Acknowledgements

We thank all the study participants and supporting personnel for their help and interest in this study. In addition, we thank the staff of the Biochemistry and Neuropsychology laboratories who collected the specimens and performed the clinical tests.

## Author contributions

MH and TY were responsible for the study conceptualization and design, drafting of the manuscript, and all statistical analyses. YM, HO, AH, YN, ME, KY, HK, ST, and MI collected and interpreted the data. BN, SY, and KE conducted the model building, training, and testing. All authors provided critical feedback regarding the analysis and the manuscript. All the authors have read and approved the final version of this manuscript.

## Availability of data and materials

All data generated or analyzed in this study are available from the corresponding author upon reasonable request, following additional ethical approval regarding data provision to individual institutions.

